# The Predictors of COVID-19 Case Fatalities in Nigerian Health Systems: A Secondary Data Analysis

**DOI:** 10.1101/2024.06.01.24308316

**Authors:** Adewale Akinjeji, Remi Oladigbolu, Adetunji Adedokun, Ogonna Onuorah, Franklin Emerenini

## Abstract

**Background:** COVID-19, caused by the novel SARS-CoV-2 is the worst catastrophe in this century that affected more than 800 million people and caused more than 7 million deaths. During the pandemic, the burden of COVID-19 increased significantly, posing a threat to public health infrastructure, testing protocols, national healthcare capacity, and disease control measures. To assess the impacts of the Nigerian Health Systems on COVID-19 fatalities, the researchers evaluated the association between healthcare system capability and mortality rate of COVID-19 patients through adjustments for healthcare spending as a proportion of the GDP, population density, and the proportion of the population that are 65 years and above across the 36 States and Abuja, FCT.

**Methods:** The study utilized secondary data abstracted from the World Bank records, Worldometer, and Post-Pandemic Health Financing by the States in Nigeria (2020 to 2022). It used data from the 36 States of the country and the FCT, Abuja. The dependent variable was COVID-19 case fatality (Case Fatality Rate across the study areas), the predictor variable was Healthcare Capacity Index (aggregate of number of doctors/nurses/midwives/hospital bed space per 1,000 population categorized into low, middle, and high Healthcare Capacity index), and the covariates were population density, health expenditure as a percentage of GDP, and the proportion of the population that are 65 years and above. A negative binomial regression model was used to assess the predictors of case fatality after adjusting for other covariates at an alpha of <0.05 and 95% confidence interval.

**Results:** Almost half of the States in Nigeria were in the middle Healthcare Capacity Index 16 (43.2%) with only 7 (18.9%) in the high Healthcare Capacity Index (HCI). The regression analysis shows that HCI was a predictor of COVID-19 case fatality as the States with high HCI compared with low HCI were 9.4 times more likely to have lower COVID-19 case fatalities (AOR=0.106, p=0.063, 95% CI[0.010-1.131]), and those with middle HCI compared with low were 6.4 times more likely to have lower COVID-19 case fatality (aOR=0.156, p=0.006, 95% CI [0.041-0.593]). Although States with a higher proportion of the population that were 65 years and above were about 2 times more likely to have higher COVID-19 case fatality (aOR 1.99, p=0.154, 95% CI [0.771-5.172]), this was not statistically significant due to the small sample size (37 States)

**Conclusion:** The research further buttressed the pivotal role that effective multidimensional healthcare capacity is a pertinent strategy to mitigate future case fatalities from Public Health Events of International Concerns (PHEICs).

## Introduction

The global impact of the COVID-19 pandemic, caused by the novel SARS-CoV-2 virus, has been profound. It has affected over 800 million people and tragically led to more than 7 million deaths worldwide(1–3). This crisis has resulted in widespread economic disruption and significant strain on healthcare systems globally, prompting unprecedented changes in social behavior, work patterns, and international cooperation that will likely shape our world for years to come(4–6).

The predictors of COVID-19 case fatalities are multifaceted and include epidemiological, demographic, and healthcare-related variables. Globally, the study by Nuhu and colleagues revealed increasing Population Median Age, higher inequalities in human development, and low vaccination rates are predictive of higher fatalities from COVID-19(7). Furthermore, the spread of COVID-19 and its associated fatalities has been linked to factors such as overcrowding, health expenditure, HIV infections, air pollution, BCG vaccination, poverty, and aging(8).

In Africa, the trends in case-fatality rates of COVID-19 revealed variations across different countries, indicating the influence of local factors on disease outcomes(9). studies revealed that the proportion of the elderly population (65 years or above) was predictive of COVID-19 fatalities(10).Other predictors of COVID-19 case fatality include population density, poverty, and urban population proportion(11).

Low and middle-income countries have borne a substantial burden of the pandemic’s impact, particularly affecting children and adolescent populations(12). Its effects have reverberated across various sectors including healthcare, the economy, education, and social dynamics worldwide(13,14). The strain on medical facilities in lower and middle-income countries like Nigeria has heightened the challenges posed by the pandemic(6). These findings highlight a crucial requirement for targeted interventions and investment in healthcare infrastructure to support vulnerable populations living in poverty or disadvantaged communities who face higher infection rates and mortality. Additionally, it emphasizes the vital role of adequate healthcare capacity in mitigating the impacts of the virus.

In Nigeria specifically, significant disparities within its health system have been revealed by this pandemic. Despite efforts made to address COVID-19’s impact, there are clear obstacles related to providing quality health services due to shortages of hospital beds, ventilators, and medical supplies which hampers citizens’ access to care during this time of crisis(15,16). Furthermore, Nigeria faces several unique challenges in combating COVID-19. These challenges include a high proportion of the population aged 65 years and above, high population density in urban areas, limited healthcare expenditure, and inadequate healthcare workforce(17). These factors contribute to the vulnerability of Nigeria’s healthcare system and its ability to effectively respond to and manage the COVID-19 pandemic. Considering the COVID-19 pandemic, Nigeria’s already fragile healthcare system faced significant challenges.

These limited resources exacerbate difficulties presented by illness leading to increased mortality rates while certain demographics especially those aged 65+ are more susceptible - sparking further examination into factors contributing towards fatality rates and how budgeting can influence outcomes within Nigeria’s healthcare infrastructure.

## Methods

Nigeria has an estimated population of 200 million people and is comprised of thirty-six states along with the Federal Capital Territory which houses Abuja, the capital city. Data on the combined total of reported COVID-19 infections and fatalities for the states in Nigeria were extracted from Worldometer, Nigeria Centre for Disease Control (NCDC) 2022 (18). The number of nurses and midwives per 1,000 people, the number of physicians per 1,000 people, and the number of hospital beds per 1,000 people were included in the analysis [Worldometer, Nigeria Centre for Disease Control (NCDC) 2022] (19).

The current healthcare costs relative to GDP, population density in terms of individuals per square kilometer (km) of land area, and the percentage of the population aged 65 or older were obtained from ONE Health finance report and World Bank data (20). In this study, we included COVID-19 cases and fatalities reported across Nigerian states, including the FCT Abuja (n=37). A composite healthcare capacity index (HCI) was computed by aggregating the counts of doctors, nurses, midwives, and available bed space, subsequently categorized into “low,” “middle,” and “high” HCI groups(21). Descriptive statistics were computed for all quantitative variables. We employed a Negative Binomial (NB) regression model to examine the relationship between outcomes and predictors/covariates. The dependent variable represented the proportion of confirmed COVID-19 cases that result in death due to the disease—referred to as case fatality rate or Incidence Rate Ratio. The primary predictor was the healthcare capacity index, while covariates encompassed population density, the proportion of individuals aged 65 or older, and health expenditure as a percentage of Gross Domestic Product (GDP). Data analysis was conducted using IBM statistical software version 25, with significance set at α < 0.05 and a confidence interval of 95%.

## Results

The median number of confirmed cases, deaths, and COVID-19 Case Fatality Rate for the 37 States in Nigeria during the review period (2022) was 2,691, 38, and 1.9. The median population density per square km was 718.9, and the median percentage of the population aged 65 and above was 2.9. The median current health expenditure as a percentage of GDP was 8.9. Furthermore, the median number of physicians, nurses and midwives, and hospital beds per 1,000 people was 0.09, 0.48, 0.05, and 0.27 respectively. Moreover, the median Healthcare capacity index was 0.89 **(Table 1)**.

**Table 1:** Descriptive statistics of selected variables (n=37).

| **Variables** | **Median** | **1<sup>st</sup> quantile to 3<sup>rd</sup> quantile** |
| --- | --- | --- |
| Case of COVID-19 | 2,691 | 1,343 – 5,620 |
| Deaths | 38 | 25 – 91 |
| Case Fatality Rate (CFR) | 1.9 | 1.1 – 2.5 |
| Population Density<sup>a</sup> | 718.9 | 672.9 – 803.7 |
| The proportion of the population ages 65 and above<sup>b</sup> | 2.9 | 2.2 – 3.3 |
| Current health expenditures | 8.9 | 6.9 – 11.3 |
| Physicians per 1000 people | 0.09 | 0.04 – 0.13 |
| Nurse and midwives per 1000 people | 0.48 | 0.26 – 0.73 |
| Midwives per 1000 people | 0.05 | 0.03 – 0.08 |
| Hospital beds per 1000 people | 0.27 | 0.17 – 0.35 |
| Healthcare Capacity Index | 0.89 | 0.50 – 1.28 |

<sup>a</sup>Population density (people per square km of land area).
<sup>b</sup>Population ages 65 and above (% of the total population).
<sup>c</sup>Current health expenditure as a percentage of GDP

**Table 2:**
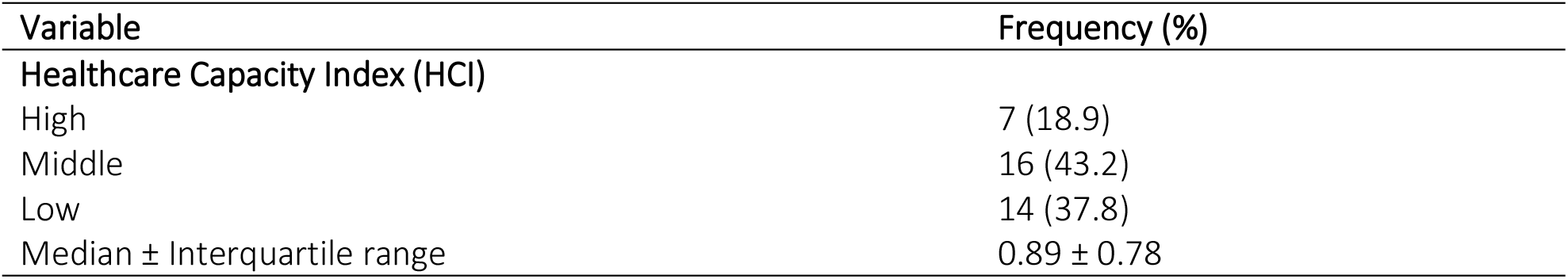
Prevalence of Healthcare Capacity Index (HCI)

**Table 2:**
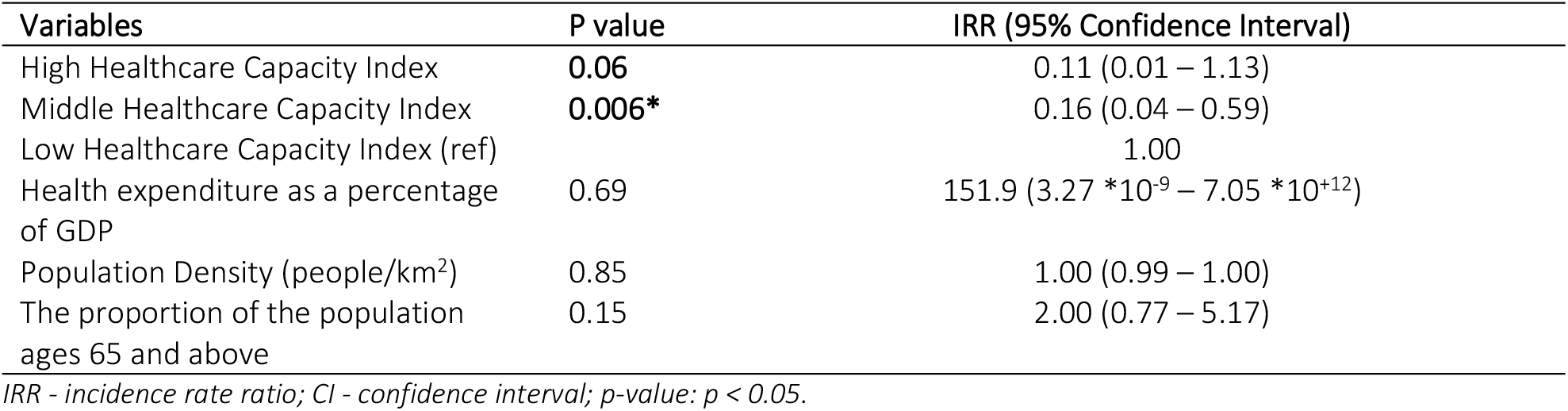
Association between Healthcare capacity and Case Fatality Rate adjusted for other variables

The Case fatality due to COVID-19 was negatively associated with the healthcare capacity index. The states with a high healthcare capacity index were 89% less likely to have high COVID-19 case fatality compared with those with low healthcare capacity index (IRR 0.11; 95% CI 0.01 – 1.13; p =0.06), similarly, the states with middle healthcare capacity index were 84% less likely to have a high COVID-19 case fatalities compared with those with low healthcare capacity index (IIR=0.16; 95% CI 0.04 – 0.59; p = 0.006). The proportion of the population aged over 65 years was positively related to the case fatality rate (IRR=2.0; 95% CI=0.77 – 1.52; p=0.15). This implies that the states with a proportion of the population ages 65 and above were 2 times more likely to have a high COVID-19 case fatality rate, however, this was not statistically significant since the p-value was 0.15. Nonetheless, this could be due to the small sample size evident by the wide confidence interval (Table 3).

Almost half 16 (43.2%) of the states in Nigeria were in middle HCI with just 7 (18.9%) of them in high HCI.

## Discussion of findings

The results of the study showed that around half of Nigeria’s states, 16 out of 37 (43.2%), had a moderate healthcare capacity index, while only 7 (18.9%) states demonstrated a high healthcare capacity index. These findings suggest that there is considerable variation in the healthcare capacity across different states in Nigeria, which may have implications for the management of COVID-19 cases and the resulting case fatalities. This finding is consistent with previous studies that have highlighted the challenges faced by the healthcare system in Nigeria, particularly in terms of infrastructure and resources(22). These challenges, such as the low number of physicians, nurses and midwives, and hospital beds per 1,000 people, can contribute to a higher case fatality rate in the context of COVID-19.

This study also reported a strong negative association between COVID-19 case fatalities and the healthcare capacity index. The findings reveal that states with a high healthcare capacity index were 89% less likely to have high COVID-19 case fatality compared to those with a low healthcare capacity index. Moreover, states with a moderate healthcare capacity index were found to be 84% less likely to experience high COVID-19 case fatalities when compared to those with a low healthcare capacity index. These significant findings align closely with previous studies, which have consistently highlighted the positive correlation between healthcare capacity and improved outcomes in effectively managing infectious diseases(23,24). Furthermore, these results support the idea that a strong healthcare system is crucial for addressing public health challenges. The availability of healthcare resources, such as hospital beds and medical supplies, plays a vital role in managing the impact of COVID-19(25,26). This underscores the importance of having robust and well-funded healthcare systems equipped with ample resources such as hospital beds, medical personnel, and advanced equipment. Such preparedness is crucial in contributing to lower COVID-19 case fatalities and ensuring better overall public health outcomes.

Additionally, according to the study findings, there was a positive association between the population aged over 65 years and the COVID-19 case fatality rate. It was observed that as the proportion of the population ages 65 and above in the states in Nigeria increases by one percentage point, they were found to be 2 times more likely to have a high COVID-19 case fatality rate, although this association was not statistically significant. These results align with previous studies that have also identified older age as a risk factor for higher COVID-19 case fatality rates in various populations around the world(27–29). These findings suggest that older individuals may be more vulnerable to severe outcomes of COVID-19, emphasizing the need for targeted efforts to protect and support this population group during the pandemic.

In conclusion, the predictors of COVID-19 case fatalities in the health systems in Nigeria include the healthcare capacity index, with higher capacity states experiencing lower case fatalities, and the proportion of the population aged over 65 years, which may contribute to higher case fatalities. Further research is needed to explore the specific mechanisms through which healthcare capacity and age impact COVID-19 outcomes in Nigeria’s health systems.

However, there was no substantial correlation found between population density, healthcare expenditure as a fraction of the gross domestic product, and COVID-19 case fatalities. These results suggest that population density and healthcare spending may not have a direct impact on COVID-19 case fatalities within Nigeria’s health systems(20,31). This underscores the necessity for a holistic strategy that considers various elements such as healthcare capacity and demographic characteristics when formulating measures to minimize the repercussions of COVID-19 and lessen case fatalities in Nigeria’s health systems. Overall, the predictors of COVID-19 case fatalities in the health systems in Nigeria include the healthcare capacity index and the proportion of the population aged over 65 years, while population density and healthcare expenditure do not appear to have a direct impact on case fatalities(32).

The findings of this study highlight the importance of robust healthcare systems with sufficient resources in effectively managing and mitigating the impact of COVID-19 and other public health events of international concerns (PHEICs). The results of the study emphasize the crucial role of healthcare systems in addressing public health challenges, particularly during a pandemic like COVID-19.

## Conclusion

The determinants of COVID-19 mortality in Nigerian healthcare systems, as indicated by this research, include the capacity of the healthcare system and the percentage of individuals aged 65 and above. These elements are linked to lower mortality rates, suggesting that a robust healthcare system and a younger population can lead to improved outcomes in managing COVID-19. The study also revealed that population density and healthcare expenditure as a share of the gross domestic product do not have a significant association with COVID-19 mortality. This implies that factors other than population density and healthcare spending may play a more substantial role in determining COVID-19 outcomes within Nigeria’s health systems. Therefore, policymakers and health authorities in Nigeria must prioritize enhancing healthcare capacity and implementing measures to safeguard the elderly populace.

The demographic structure and healthcare capacity emerge as vital predictors of COVID-19 fatalities within Nigerian health systems, underlining the necessity for an all-encompassing approach to pandemic response addressing these aspects. These determinants underscore how important it is for policymakers to develop targeted interventions based on such factors when seeking ways to mitigate the impact of COVID-19 - including bolstering medical resources, instituting preventive measures targeting older adults, and focusing on vaccination initiatives. Moreover, addressing shortages in the medical workforce while improving infrastructure are critical steps toward reinforcing the ability of Nigeria’s health system to respond effectively during public health crises like COVID-19.

The study used secondary data, which may have limitations in accuracy and completeness. Also, it focused specifically on Nigeria’s healthcare system and may not be generalizable to other countries or regions. Additionally, the study only examined a limited number of predictors of COVID-19 fatalities and did not consider other potential factors such as socioeconomic status, access to healthcare services, and availability of critical medical equipment.

## Data Availability

World Bank records (https://data.worldbank.org/indicator/SH.MED.NUMW.P3?locations=NG), Worldometer (https://www.worldometers.info/), and Post-Pandemic Health Financing by the States in Nigeria 2020 to 2022 (https://cdn.one.org/pdfs/ONE_2022_Nigeria_Budgetary_Health_Report.pdf)

https://data.worldbank.org/indicator/SH.MED.NUMW.P3?locations=NG

